# Non-Invasive Mechanical Ventilation reduces the Motor Decline in Amyotrophic Lateral Sclerosis

**DOI:** 10.1101/2023.05.26.23290514

**Authors:** Maurizio Grassano, Emanuele Koumantakis, Umberto Manera, Antonio Canosa, Rosario Vasta, Palumbo Francesca, Giuseppe Fuda, Paolina Salamone, Giulia Marchese, Federico Casale, Lorena Charrier, Gabriele Mora, Cristina Moglia, Andrea Calvo, Adriano Chiò

**Author notes:** **Corresponding author:** Maurizio Grassano, ALS Center, “Rita Levi Montalcini” Department of Neuroscience, University of Turin, Via Cherasco 15, Turin, Italy, 10126. These authors contributed equally to this work (Andrea Calvo, Adriano Chiò).

## Abstract

**Background:** Non-invasive mechanical ventilation (NIMV) improves Amyotrophic Lateral Sclerosis (ALS) quality of life and survival. However, data about its effect on disease progression are still lacking.

**Objective:** To test whether NIMV use changed the rate of functional decline among ALS patients.

**Methods:** In this retrospective observational study, we included 465 ALS patients followed up at the ALS Center in Turin, Italy, who underwent NIMV during the disease course. The primary outcome was the change in functional decline after NIMV initiation when adjusting for covariates. Functional decline was based on the non-respiratory items of the ALS Functional Rating Score – Revised (ALSFRS-R).

**Results:** A slower progression of functional decline followed NIMV initiation (mean improvement 0.13, 95%CI 0.11 to 0.16, p<0.001) regardless of sex, age at diagnosis, and disease duration prior to NIMV initiation. Indeed, the disease stage at NIMV initiation (early or advanced) did not influence the results. Respiratory support exerts its slowing effect mainly on the progression of spinal motor function.

**Conclusions:** We proved that NIMV influences the rate of motor progression in ALS. This result was not a consequence of the ALSFRS-R floor effect. The functional decline slowed after starting NIMV independently of the site of disease onset. Our results reinforce the importance of not delaying NIMV initiation in all ALS patients. NIMV-induced slowing of disease progression should also be accounted for when evaluating clinical trial outcomes.

## Background

Amyotrophic lateral sclerosis (ALS) is a progressive neurodegenerative disorder that causes disability and death, usually because of respiratory failure within 3 years of onset^1^. A significant advance in the management of ALS was the demonstration of the beneficial effects of non-invasive ventilation (NIMV) on patients’ survival and quality of life^2^. Therefore, the mechanical support of respiratory function is now regarded as an essential component of care^3,4,5^. Although several studies have reported that NIMV reduced the decline of pulmonary function, its effect on patients’ functional outcome and motor progression is still controversial^6^. Here, we evaluated whether respiratory support slows the progression of functional decline in a large cohort of ALS patients.

## Methods

### Study population

All patients followed-up at the ALS Center in Turin, Italy and included in the Piemonte and Valle d’Aosta Register for ALS (PARALS) from January 1, 2007, to December 31, 2019 were considered eligible. We included in the study all ALS patients who underwent NIMV and had available ALSFRS-R follow-up data after the date of NIMV initiation. The other characteristics of the PARALS have been detailed elsewhere^7^.

### Main variables and outcomes

ALSFRS-R was collected for each patient at the ALS clinic approximately every 2-3 months. For the analysis of motor function, we only considered the non-respiratory items (items 1 to 9) of the ALSFRS-R (non-respiratory ALSFRS-R). Additionally, the decline rates of ALSFRS-R subdomains were evaluated as secondary outcomes: the bulbar ALSFRS-R was derived from items 1 to 3 of the ALSFRS-R scale; the motor ALSFRS-R was calculated from items 4 to 9 (eMethods). Diagnosis of frontotemporal cognitive and behavioral syndromes was based on cognitive assessments and consensus criteria^8^. Additionally, King’s^9^ and MiToS^10^ stages were calculated from ALSFRS-R scores (eMethods). Clinical stage at the visit before NIMV initiation was used to determine whether the ventilatory support benefits patients across different functional stages of ALS. We defined early disease stages as MiToS stage 0 and King’s stage 0 or 1; MiToS stages ≥ 1 and King’s stages ≥ 2 were instead considered advanced disease stages. Age at symptoms onset, sex, presence of cognitive impairment, and site of disease onset were used as additional covariates.

### Statistical methods

To analyze longitudinal progression and changes of the functional decline in ALS patients, we implemented univariate and multivariable Mixed-Effects Regression Models (MERs), with patients’ effect fitted as random. MERs advantages over fixed-effects models are detailed in eMethods. Additionally, we performed a sensitivity analysis using the generalized least square (GLS) regression, to assure the replicability of the results adopting a different statistical methodology (eMethods, eTable 1).

### Ethics approval

This study was approved by the Comitato Etico Azienda Ospedaliero-Universitaria Città della Salute e della Scienza, Torino (#355732). All participants gave informed consent.

### Data availability

Data is available to interested researchers upon motivated and reasonable request.

## Results

A total of 465 patients were included in this analysis. Mean age at disease diagnosis (SD) was 66.5 years (10.2), with 28.6% of patients with bulbar onset and 39.2% of patients with cognitive impairment. Patients initiated NIMV after a median (IQR) 8.94 months (2.33 – 18.6). Baseline demographic and disease characteristics of the patients’ cohort are summarized in eTable 2.

### Effect of NIMV on disease progression

Longitudinal analysis revealed that initiating ventilatory support caused a deceleration of disease progression compared to the pre-NIMV period as measured by non-respiratory ALSFRS-R (mean improvement 0.13, 95%CI 0.10 to 0.16, p< 0.001, eTable 3). This monthly slowing remained significant in the multivariable analysis (mean improvement 0.13, 95%CI 0.11 to 0.16, p< 0.001) (Figure 1, Figure 2).

**Figure 1.**
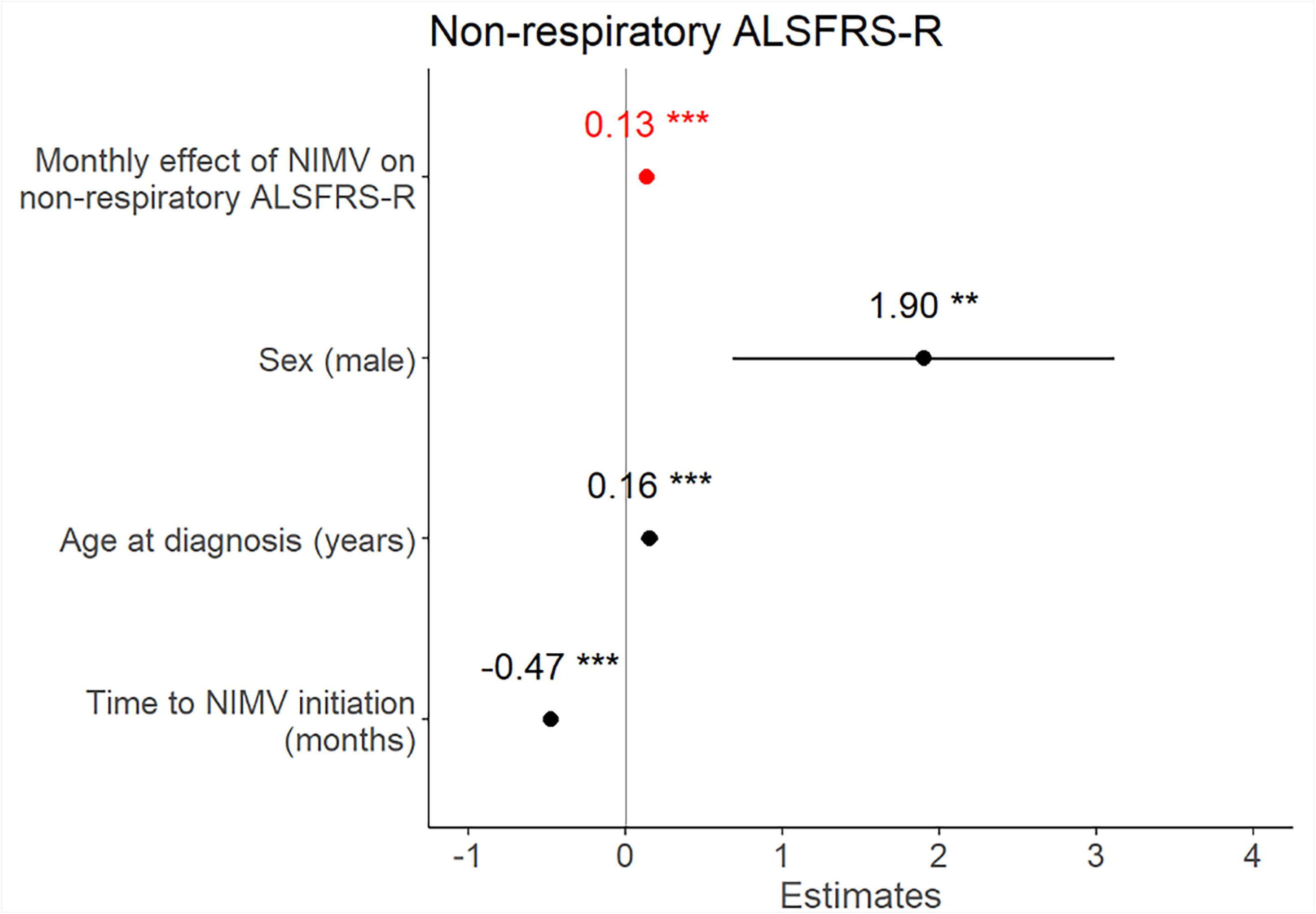
Forest plot summarizing multivariable linear mixed-effects model’s fixed effects on non-respiratory ALSFRS-R. The main independent variable was NIMV-related monthly ALSFRS-R change, adjusted for sex, age at diagnosis, time to/from NIMV initiation. *** p≤0.001, ** p≤0.01, *p≤0.05

**Figure 2.**
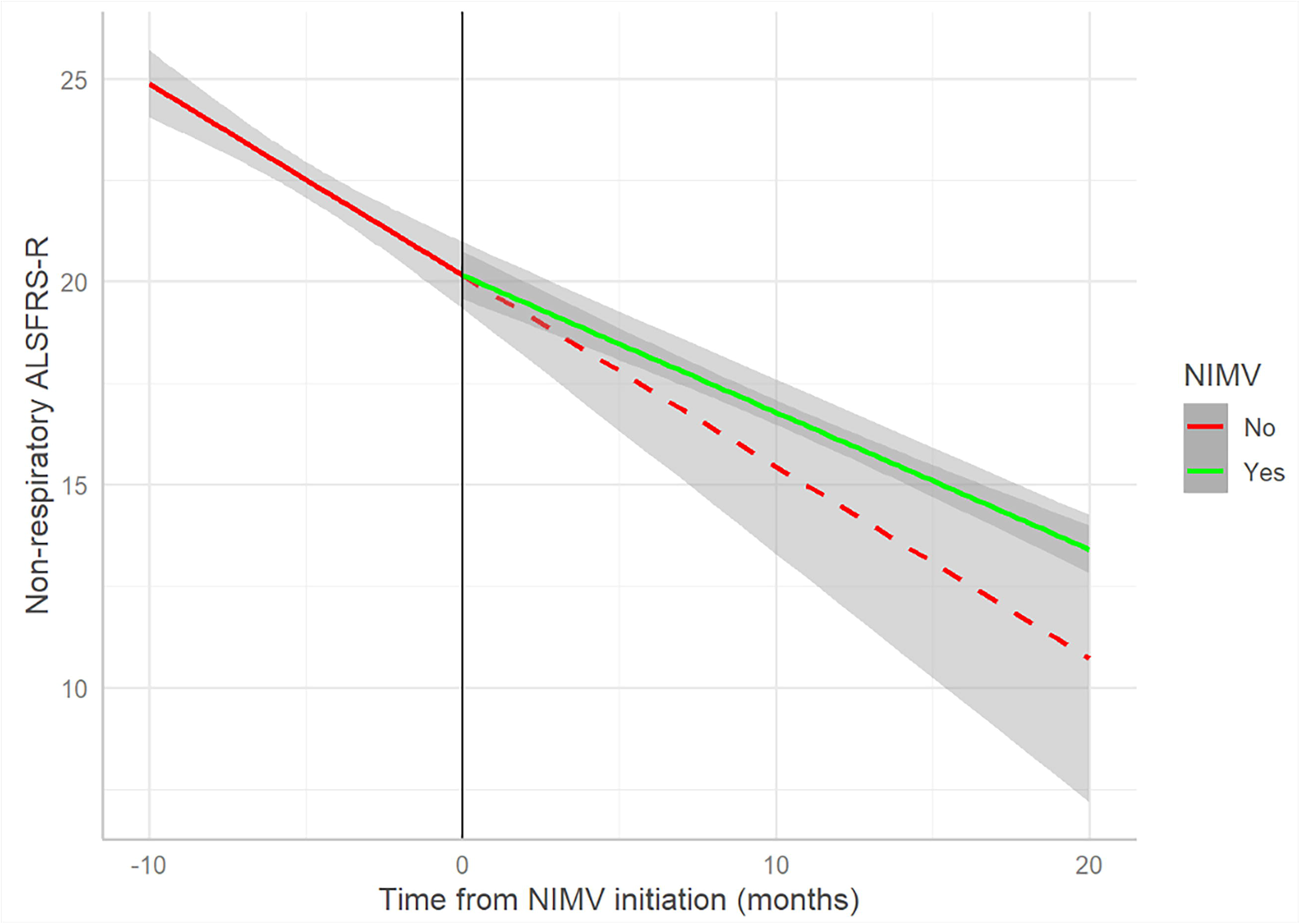
Multivariable linear mixed-effects model’s predicted values of non-respiratory ALSFRS-R according to time from NIMV initiation. The red line represents the predicted values based on the scores of the patients before NIMV initiation, while the green line represents the predicted values based on the observations after the start of NIMV. The dashed red line was also extended for time points after NIMV initiation, representing the score prediction in the case the patients had not started NIMV. Confidence intervals (grey area) were reported for both curves. The difference in slope between the two prediction lines is representative of the monthly effect of NIMV on disease progression.

### Effect of NIMV according to disease stage

We performed additional analysis adjusting for disease stage at the time of NIMV initiation: the results revealed that NIMV usage reduced the rate of ALS progression regardless of disease severity at the time of NIMV as evaluated by either MiToS or King’s (mean improvement 0.13, 95%CI 0.10 to 0.16, p< 0.001) (eFigure 1).

### NIMV effects on bulbar and spinal function

Analysis using spinal and bulbar ALSFRS-R subdomains showed that NIMV determined a prominent deceleration in motor functions decline (mean improvement 0.13, 95%CI 0.11 to 0.16, p< 0.001), while it did not show a significant effect on bulbar symptoms’ progression (mean 0.00, 95%CI -0.01 to 0.01, p= 0.956) (eFigure 2). However, both patients with spinal and bulbar disease onset experienced the NIMV-related progression slowing (eFigure 3).

**Figure 3.**
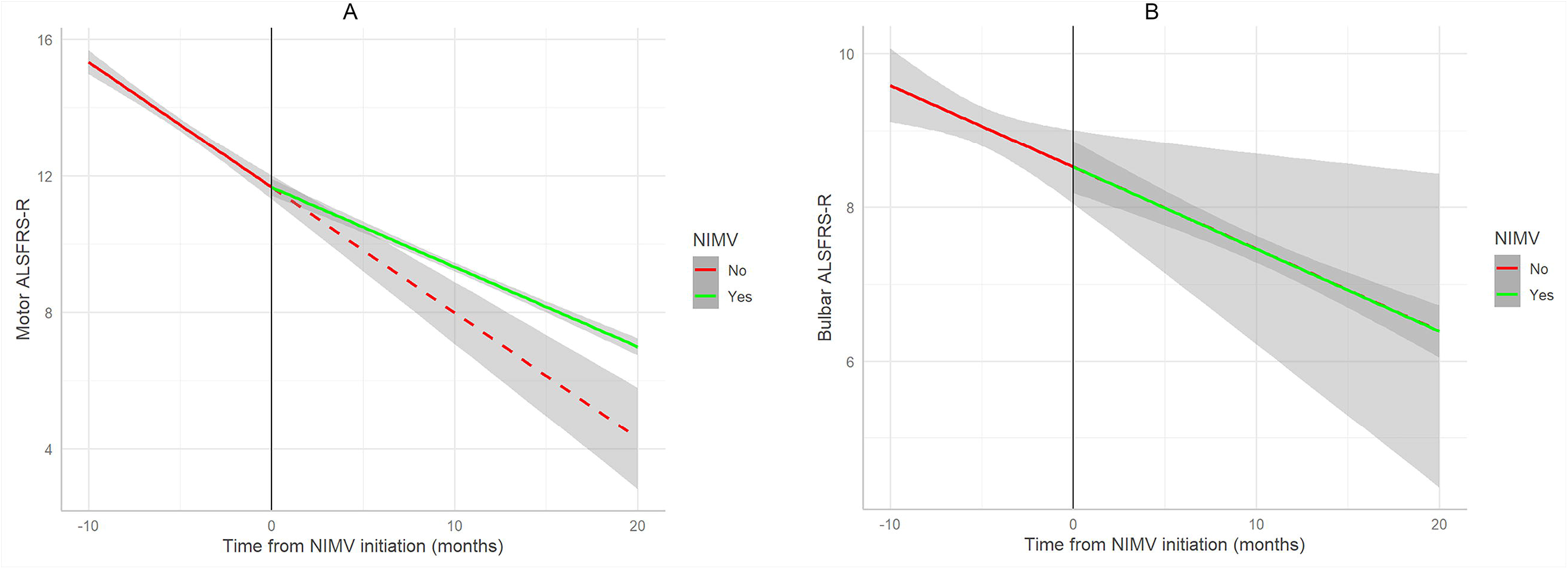
Multivariable linear mixed-effects model’s predicted values of motor (A) and bulbar (B) ALSFRS-R according to time from NIMV initiation. The red line represents the predicted values based on the scores of the patients before NIMV, while the green line represents the predicted values based on the observations after the start of NIMV. The dashed red line was also extended for time points after NIMV initiation, representing the score prediction in the case the patients had not started NIMV. Confidence intervals (grey area) were reported for both curves. The difference in slope between the two prediction lines is representative of the monthly effect of NIMV on disease progression.

## Discussion

We evaluated the change in functional decline after NIMV initiation in a large Italian ALS cohort and observed that NIMV reduced the motor decline measured by the ALSFRS-R scale. To the best of our knowledge, this is the first study to demonstrate that mechanical ventilation slows ALS motor progression as measured with ALSFRS-R (Figure 2).^6^ This positive NIMV-related effect was observed independently of the disease stage, thus suggesting that initiating NIMV earlier during the disease course could also significantly benefit patients’ functional decline. NIMV especially helped preserve motor functions (Figure 3) with a more pronounced effect on bulbar onset patients (eFigure 3). Together, our data suggest that NIMV benefits all ALS patients, independently of age, sex, time to NIMV initiation, site of disease onset, and cognitive function (eTable 4).

Our findings add to the well-known effects of NIMV on patients’ survival and quality of life^2^. In addition, they have relevant clinical implications. First, because NIMV slows the rate of motor decline, it is increasingly crucial to perform regular respiratory evaluations even in patients without overt respiratory symptoms and initiate NIMV without delay when criteria are met.

Moreover, discussing with patients and their caregivers this direct disease-slowing effect may help to improve their adherence to mechanical respiratory support. Third, our findings should be carefully considered in trial designs and analyses, where disease progression is a major endpoint outcome^11^. Indeed, since our models demonstrated that the positive impact of respiratory support is immediate, assessing NIMV as a potential confounder should be recommended.

The positive effect of NIMV on survival and quality of life has been attributed to the improvement of hypoventilation and the relief of dyspnea^12^. There is currently no precise putative mechanism explaining how NIMV could affect motor function. It has been proposed that NIMV usage reduces respiratory-related energy expenditure. Decreasing the work of breathing and its consequent caloric burn could improve weight loss and muscle wasting due to hypermetabolism, thus in part protecting against further motor damage^13^.

Our study has some limitations. First, our results may have been affected by the intrinsic limitations in measuring ALS progression, namely the floor effect of the ALSFRS-R scale^14^ and the variable slope of ALSFRS-R trajectories. However, we employed a model that predicted the ALSFRS-R using the measurement immediately prior in time and additionally included disease staging as a covariate to further validate our findings. Second, because of the observational setting, the quantitative effect of NIMV on ALSFRS-R progression presented here is not based on a randomized controlled trial. Nevertheless, our large cohort with prospectively collected observational data may represent more realistic conditions and provide more generalizable real-world data.

In conclusion, our longitudinal model and sensitivity analyses provide robust and substantial evidence for a causal effect of NIMV on disease slowing and improved survival. Optimizing respiratory support is a significant step toward effective care for ALS.

## Data Availability

Data is available to interested researchers upon motivated and reasonable request.

## Contributorship Statement

Maurizio Grassano: study concept and design; data analysis; drafting of the manuscript; critical revision of the manuscript for important intellectual content. Emanuele Koumantakis: data analysis; drafting of the manuscript; critical revision of the manuscript for important intellectual content. Umberto Manera: data collection; critical revision of the manuscript for important intellectual content. Antonio Canosa: data collection; critical revision of the manuscript for important intellectual content. Rosario Vasta: data collection; critical revision of the manuscript for important intellectual content. Palumbo Francesca: data collection; critical revision of the manuscript for important intellectual content. Giuseppe Fuda: data collection; critical revision of the manuscript for important intellectual content; administrative, technical and material support. Paolina Salamone: data collection; critical revision of the manuscript for important intellectual content; administrative, technical and material support. Giulia Marchese: data collection; critical revision of the manuscript for important intellectual content; administrative, technical and material support. Federico Casale: data collection; critical revision of the manuscript for important intellectual content; administrative, technical and material support. Lorena Charrier: data analysis; drafting of the manuscript; critical revision of the manuscript for important intellectual content. Gabriele Mora: study concept and design; drafting of the manuscript; critical revision of the manuscript for important intellectual content; study supervision. Cristina Moglia: study concept and design; drafting of the manuscript; critical revision of the manuscript for important intellectual content; study supervision. A Calvo: study concept and design; drafting of the manuscript; critical revision of the manuscript for important intellectual content; study supervision. Adriano Chiò: study concept and design; drafting of the manuscript; critical revision of the manuscript for important intellectual content; obtained funding; study supervision.

