## Supplementary File for "Non-Invasive Mechanical Ventilation reduces the Motor Decline in Amyotrophic Lateral Sclerosis"

**Table of contents**

**eMethods	2-3**

**eTable 1.** Generalized Least Squares Model	**4**

**eTable 2.** Baseline characteristics of the study cohort	**5**

**eTable 3.** Univariate Linear Mixed-Effects Model (non-respiratory ALSFRS-R)	**6**

**eTable 4.** Multivariable Linear Mixed-Effects Model (non-respiratory ALSFRS-R)	**7**

**eFigure 1.** Multivariable Linear Mixed-Effects Model’ Forest Plot (non-respiratory

ALSFRS-R), adjustment for MiToS and King’s stage	**8**

**eFigure 2.** Multivariable Linear Mixed-Effects Model’ Forest Plot (motor or bulbar

ALSFRS-R)	**9**

**eFigure 3.** Multivariable Linear Mixed-Effects Model’ Forest Plot (non-respiratory

ALSFRS-R), stratification for disease site of onset (spinal or bulbar)	**10**

**eTable 5.** Multivariable Linear Mixed-Effects Model (non-respiratory ALSFRS-R),

adjustment for cognitive impairment	**11**

#### eMethods

**Piemonte and Valle d’Aosta Register for ALS (PARALS)**

The PARALS is a prospective register of all cases of ALS in the Piemonte and Valle d’Aosta regions of Italy, whose epidemiologic data regarding the 1995-2004 and 2005-2014 periods have previously been published. The register database is anonymized and treated according to the Italian Data Protection Code. The primary sources of cases are the two tertiary ALS centers, located in Torino and Novara, and the neurology departments of the two regions. The diagnosis of ALS was made after evaluating relevant clinical information and based on the revised El Escorial Criteria (EEC-R). Patients are included in the PARALS if they meet the diagnosis of definite, probable, or probable laboratory-supported ALS according to the EEC-R at any stage of the disease. Follow-up visits of each patient are performed at regular intervals (2-4 months), and ad hoc questionnaires are administered to collect data on disease history, ALS functional rating scale and treatments.

**Criteria for NIV initiation**.

The indication for NIMV was based on current guidelines ^1,2^. Briefly, patients initiated NIMV after pneumological evaluation and if: vital capacity (VC) was <80% to <50% of predicted values; maximum inspiratory pressure (MIP) was <40 cm to < 60 cm H_2_O; sudden nasal inspiratory pressure (SNIP) was <40 cm H_2_O; in nocturnal pulse oximetry the % of time spent <90%, was >5% or >10%; in arterial blood gas analysis the PCO_2_ was > 45 mm Hg.

**Disease staging**

King’s staging is based on the spreading of motor symptoms in three different body regions (bulbar, upper limbs, and lower limbs) and on the use of non-invasive ventilation (NIV) and enteral nutrition. The bulbar region was considered involved if a patient lost any points on any of the three items regarding speech, salivation, or swallowing (items 1, 2, and 3). The upper limb region was considered involved if a patient lost any points on either items 4 and 5A (handwriting and ability to cut food and handle utensils). The lower limb region was considered involved if a patient lost any points on the item regarding walking (item 8). The presence of gastrostomy was confirmed by the assessment of item 5B (evaluation of the ability to manipulate fastenings if a patient has a gastrostomy) rather than item 5A (answered by patients without gastrostomy). If a subject scored 0 points on question 10 (indicating that the patient has significant difficulty with dyspnea and is considering using mechanical respiratory support) or less than 4 points on question 12 (dropping any points on this question indicates that Bi-level airway pressure ventilation is being used), this indicated that the patient was using NIV. We classified patients according to the following five stages of King’s staging system: 1, one region involved; 2, two regions involved; 3, three regions involved; 4A, patient needs gastrostomy; 4B, patient needs NIV.

MiToS staging of ALS progression is defined by loss of independent function in four key domains that are included in ALSFRS-R: walking/self-care, swallowing, communicating and breathing. Impairment in each domain was determined by thresholds that reflected loss of autonomy in the specific ALSFRS-R. Movement was considered impaired if the score of item 6 (dressing and hygiene) or item 8 (walking) was below 2; communication was considered impaired if patient lost more than two points in both item 1 (speech) and 4 (handwriting); swallowing was considered impaired if patient needed supplemental tube feeding or exclusively parenteral or enteral feeding on item 3 (swallowing); breathing was considered impaired if the score of either item 10 (dyspnea) or 12 (respiratory insufficiency) was below 2. Values of 0 (below threshold) or 1 (above threshold) were assigned, and the stages were determined as the sum of those values across the four domains. Stages were defined as follows: stage 0, functional involvement but no loss of independence on any domain; stages 1–4, number of domains in which independence was lost; and stage 5, death.

**Cognitive impairment.**

ALS patients underwent an extensive cognitive battery at diagnosis^3^ and were classified according to the consensus criteria for the diagnosis of frontotemporal cognitive and behavioral syndromes in ALS^4^. Cognitive status was included as a covariate because behavioral or cognitive dysfunction in ALS has been demonstrated to have a negative effect on NIV usage and disease outcome. For the purpose of our analysis, we confronted cognitive normal patients with ALS patients with cognitive impairment. Since we included in the study ALS patients from January 1, 2007, to December 31, 2019, neuropsychological assessment was available only for a subgroup of patients: therefore, analysis with cognitive status as a covariate were performed separately and reported in Supplementary Results.

**Statistical Analysis**

We fitted Mixed-Effect Regression models (MERs), considering non-respiratory ALSFRS-R score as outcome variable and considering patients’ effect as random. MERs are advantageous over fixed-effects models in that (i) they capture correlations of repeated measures using “random effects” that describe specific trends over time, therefore not assuming that all observations are independent (fitting a covariance pattern). (ii) When dealing with repeated measures, MERs do not require complete data from all subjects, unlike fixed-effects models. This results in more appropriate estimates of the effect of treatment and their standard errors (SEs). In this line, MER is more robust to missing data, provided that it can be assumed missing at random. (iii) MERs can estimate overall treatment effects and at the same time estimate treatment effects at each time point. Standard errors for treatment effects at individual time points are calculated using information from all time points and are therefore more robust than standard errors calculated from separate time points^5^.

Univariate analysis was first performed, evaluating interaction between time from diagnosis and NIMV use as main independent variable (eTable 3). Then, multivariable models were fitted, adjusting for sex, age at diagnosis (Figure 1, eTable 4), and, as secondary analyses, also MiToS o King’s Stage (eFigure 1). Moreover, multivariate analyses were performed with motor or bulbar ALSFRS-R score as dependent variable (eFigure 2). Lastly multivariable models were fitted also adjusting for cognitive impairment, which showed to maintain NIMV effect significance (eTable 5). However, as cognitive status was only evaluated in 337 (72.5%) patients, these results were not reported in the main text. As ALSFRS-R is thought to not follow a linear decrease in patients, in all the previous models we implemented a covariance pattern using a type 1 autocorrelation, which predicts the ALSFRS-R of follow-ups using the immediately prior value in time (ALSFRS-R in the previous follow-up).

As a sensitivity analysis, Generalized Least Squares (GLS) models were used (eTable 1), because of their advantages in the analysis of observational longitudinal data. GLS models can be used for data that does not follow a normal distribution, which is a requirement for traditional linear regression. Moreover, they can account for non-ignorable missing data, where the data is missing not at random and could impact the results. Additionally, they allow widely varying observations per subject and account for subjects to have distinct trajectories. Finally, they are not biased, especially in case of non-random sample dropouts^6^. However, for our main results we decided to report only the effects of MERs, as these models can accommodate non-linear relationships using random effects and can incorporate subject-specific information into the analysis, providing a more accurate understanding of the data.

Statistical analyses were carried out using R, version 4.1.2^7^. MERs were performed using the “lme” function from “nlme” R package, while GLS models used the “GLS” function in “rms” R package. All statistical tests were two-sided, with a level of statistical significance set at 0.05.

#### eResults

#### eTable 1. Generalized Least Squares (GLS) multivariable model’s estimates, with corresponding 95% confidence intervals and p-values. Outcome variable was non-respiratory ALSFRS-R, while the main independent variable was the interaction between time from/to NIMV initiation (in months) and use of NIMV, adjusted for sex and age at diagnosis.

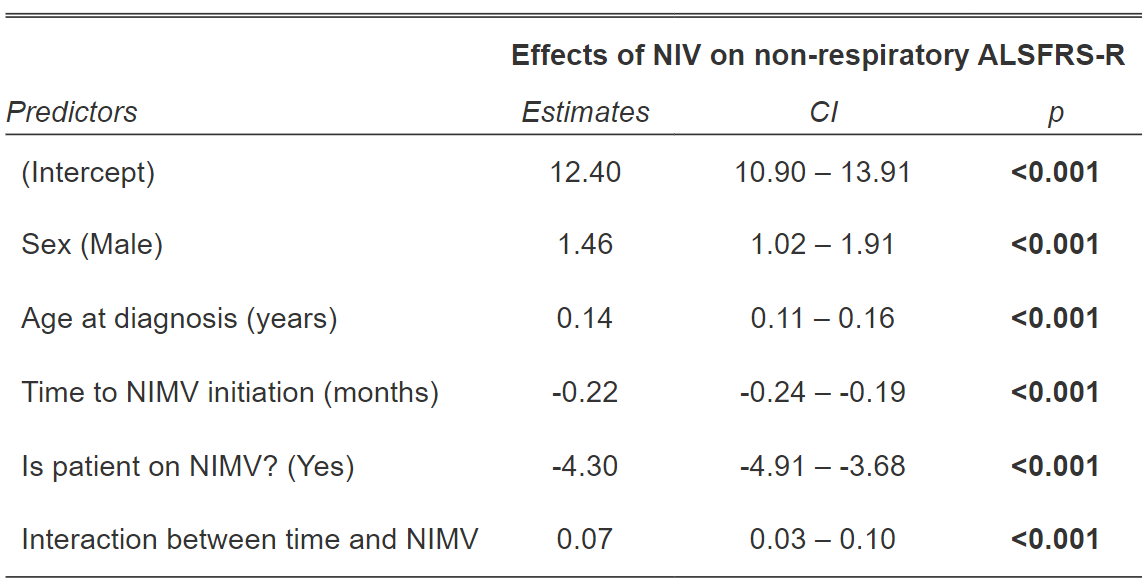

#### eTable 2. Baseline characteristics of the study cohort. From the 623 eligible ALS patients who were administered NIMV during our study period, 158 subjects were excluded from the analyses as no follow-up after NIMV initiation was available. As expected, excluded patients had shorter follow-up time and initiated NIMV later during the disease course (after the last available follow-up). Excluded and included subjects did not differ in terms of other relevant demographic or clinical variables.

|  | **Included**  **(N=465)** | **Excluded**  **(N=158)** | **p-value*** |
| --- | --- | --- | --- |
| **Sex:** Male | 279 (60.0%) | 87 (55.1%) | 0.320 |
| **Age at diagnosis (years)** |  |  |  |
| Mean (SD) | 66.5 (10.2) | 67.0 (10.6) | 0.597 |
| **Follow-up time (months)** |  |  |  |
| Median (IQR) | 17.7 (6.90-32.0) | 7.06 (0-20.6) | **<0.001** |
| **Time from diagnosis to NIMV (months)** |  |  |  |
| Median (IQR) | 8.94 (2.33-18.6) | 11.1 (4.76-21.0) | **0.038** |
| **Diagnosis delay (months)** |  |  |  |
| Median (IQR) | 0.75 (0.42-1.08) | 0.67 (0.42-1.06) | 0.429 |
| **Onset:** Bulbar | 133 (28.6%) | 49 (31.8%) | 0.511 |
| **Cognitive impairment** | 132 (39.2%) | 48 (45.7%) | 0.281 |
| **Basal FVC** |  |  |  |
| Median (IQR) | 84 (65-100) | 90 (63-103) | 0.427 |
| **ALSFRS-R at diagnosis** |  |  |  |
| Median (IQR) | 42 (37-45) | 42 (37-45) | 0.653 |
| **Medium/High MiToS stage at NIMV start** | 89 (19.1%) | 18 (13.8%) | 0.208 |
| **Medium/High King's stage at NIMV start** | 276 (59.4%) | 82 (63.1%) | 0.506 |

#### NIMV, non-invasive mechanical ventilation; SD, standard deviation; IQR, interquartile range.

#### Categorical variables are presented as number (percentage) of patients.

*Chi-squared test was performed for categorical variables; for numerical variables, t-test was used when means are reported, while Wilcoxon rank sum test was used when medians are reported.

#### eTable 3. Univariate linear mixed-effects model’s fixed effects (interaction between NIMV use and months from/to NIMV initiation) and random effects (patients’ effect) on non-respiratory ALSFRS-R. Corresponding 95% confidence intervals and p-values are reported.

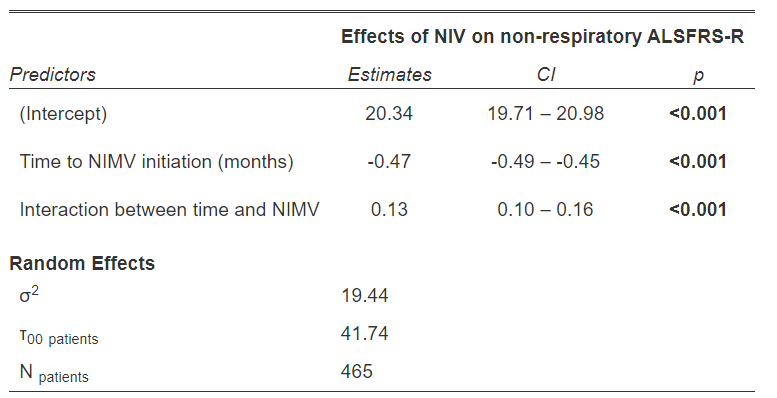

#### eTable 4. Multivariable linear mixed-effects model’s fixed effects (sex, age at diagnosis, time from/to NIMV initiation, and interaction between NIMV use and months from/to NIMV initiation) and random effects (patients’ effect) on non-respiratory ALSFRS-R. Corresponding 95% confidence intervals and p-values are reported.

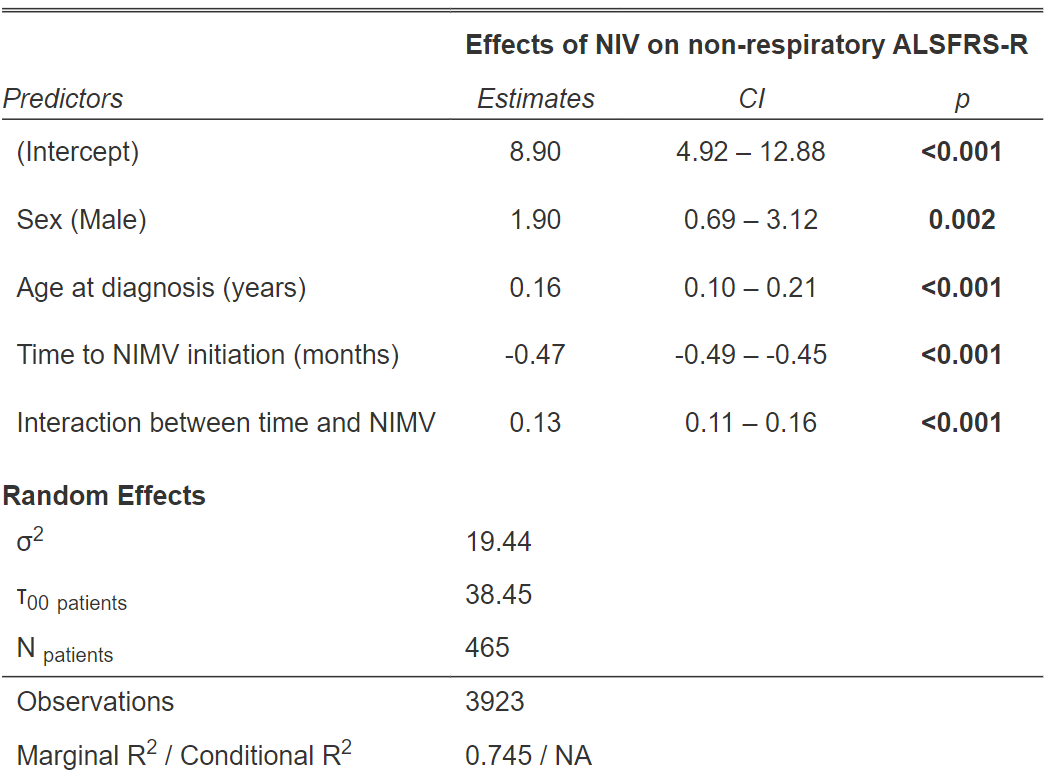

#### eFigure 1. Forest plots summarizing multivariable linear mixed-effects model’s fixed effects on non-respiratory ALSFRS-R. The main independent variable was NIMV-related monthly ALSFRS-R change, and adjustments were made for sex, age at diagnosis, time to/from NIMV initiation, and MiToS or King’s stage at NIMV initiation.

*** p≤0.001, ** p≤0.01, *p≤0.05

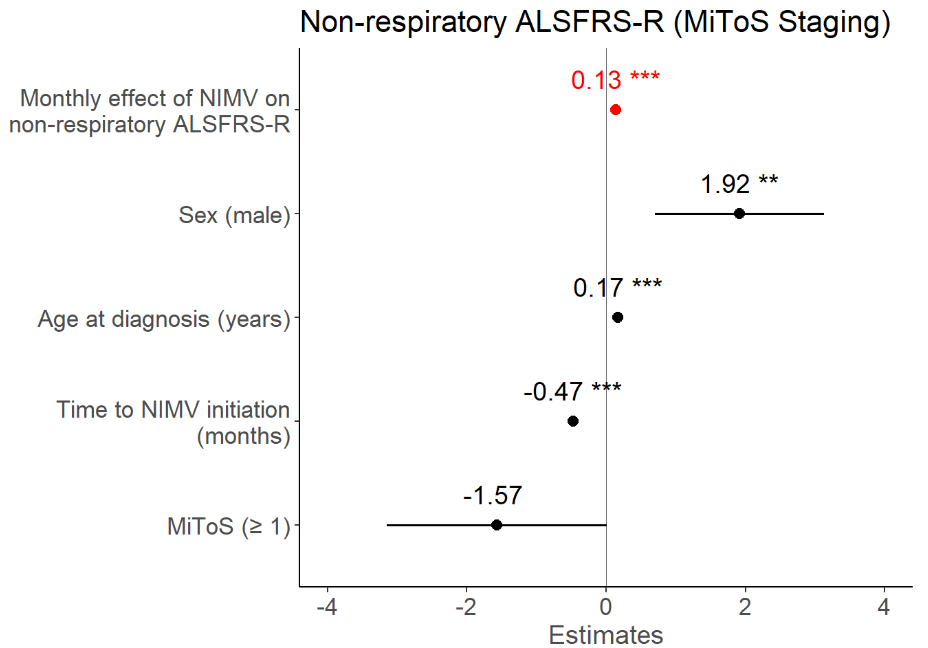

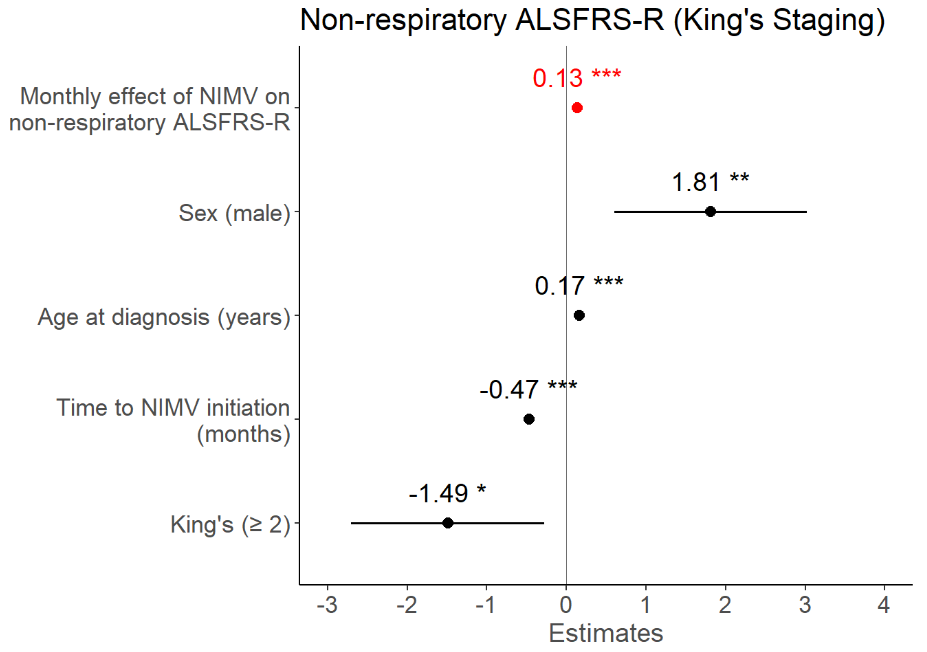

#### eFigure 2. Forest plots summarizing multivariable linear mixed-effects model’s fixed effects on motor ALSFRS-R and bulbar ALSFRS-R. The main independent variable was NIMV-related monthly ALSFRS-R change, and adjustments were made for sex, age at diagnosis, and time to/from NIMV initiation.

*** p≤0.001, ** p≤0.01, *p≤0.05

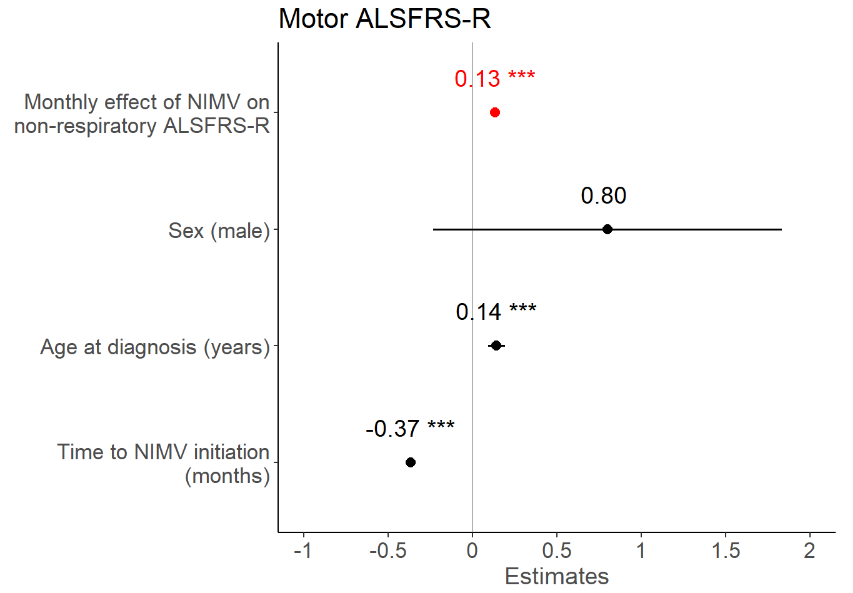

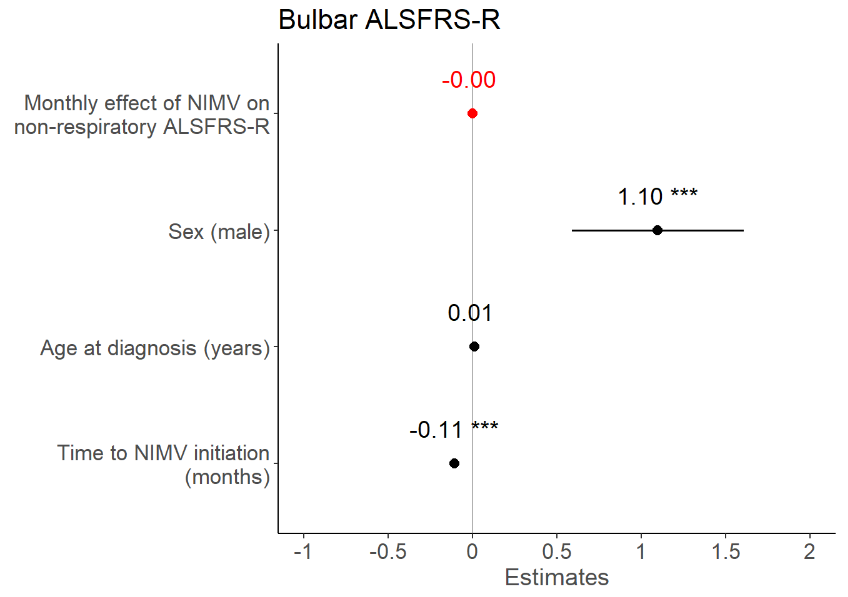

#### eFigure 3. Forest plots summarizing multivariable linear mixed-effects model’s fixed effects on non-respiratory ALSFRS-R. The main independent variable was NIMV-related monthly ALSFRS-R change, and adjustments were made for sex, age at diagnosis, and time to/from NIMV initiation. Analyses were stratified for disease site of onset (spinal or bulbar).

*** p≤0.001, ** p≤0.01, *p≤0.05

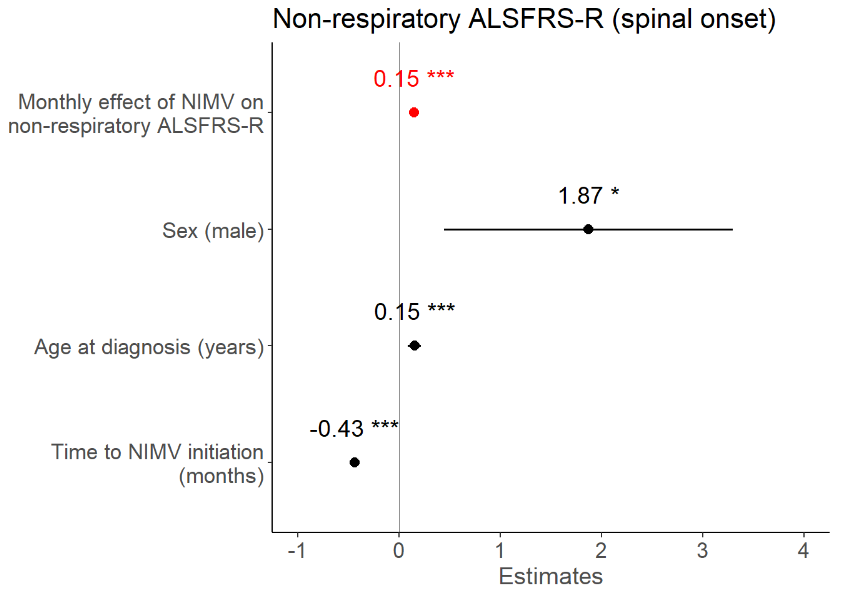

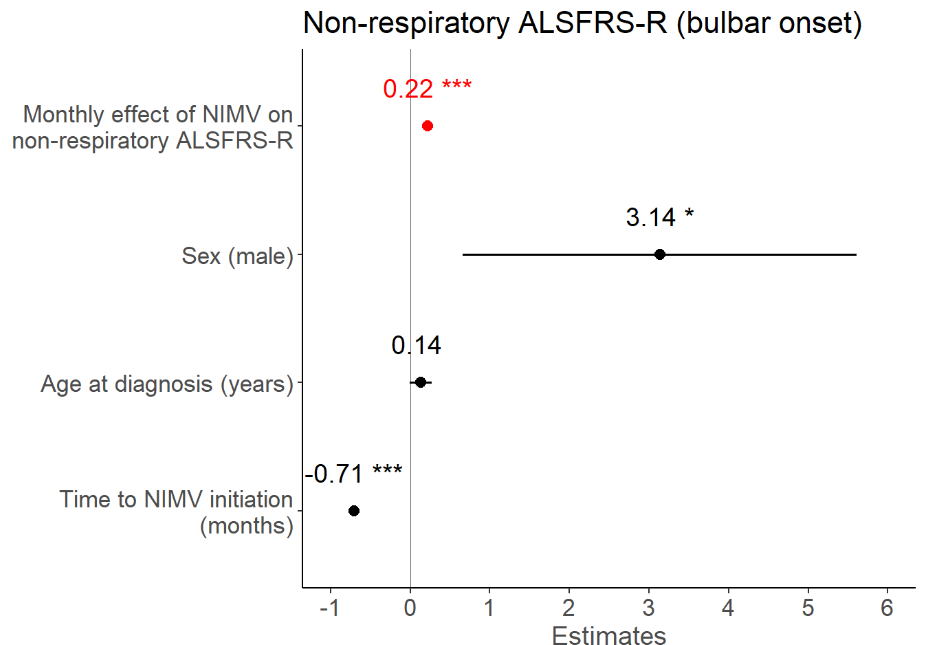

#### eTable 5. Multivariable linear mixed-effects model’s fixed effects (sex, age at diagnosis, cognitive impairment, time from/to NIMV initiation, and interaction between NIMV use and months from/to NIMV initiation) and random effects (patients’ effect) on non-respiratory ALSFRS-R. Corresponding 95% confidence intervals and p-values are reported.

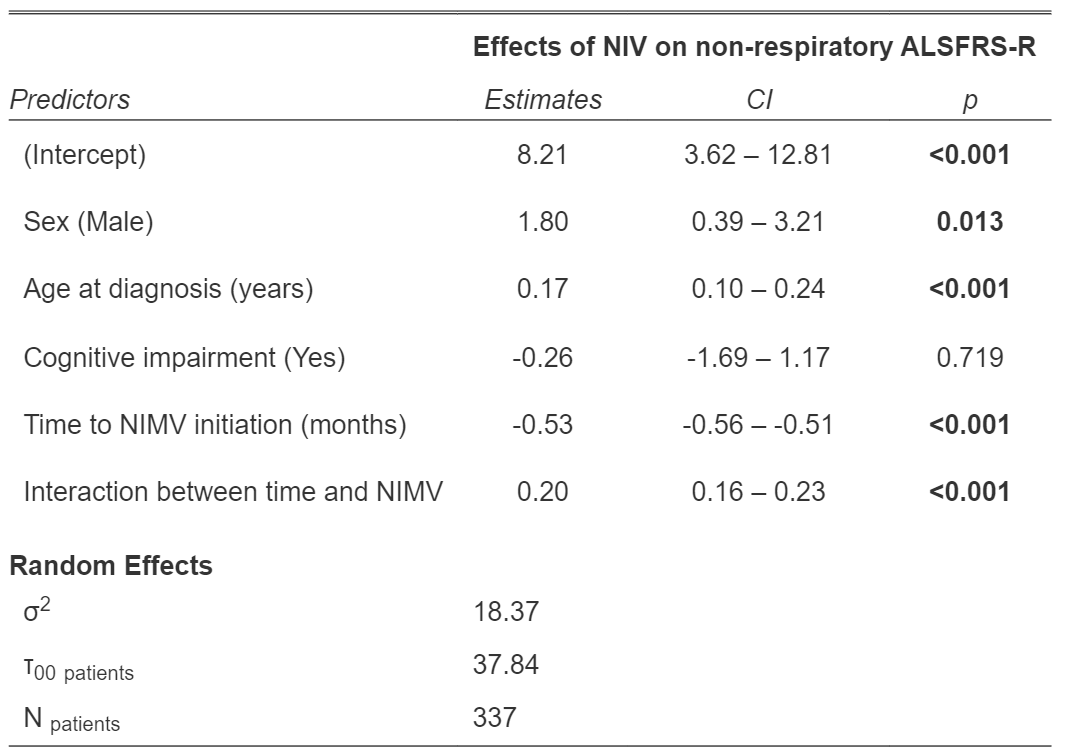
